# Dietary Assessment and Prevention of Hypertension in Nigeria: Protocol for a Retrospective Cross-sectional Study for the Development and Validation of a Food Frequency Questionnaire for Clinical Use

**DOI:** 10.1101/2023.09.25.23296109

**Authors:** Nimisoere P. Batubo, J. Bernadette Moore, Michael A. Zulyniak

**Affiliations:** Nutritional Epidemiology Group, School of Food Science and Nutrition, University of Leeds, Leeds, LS2 9JT

## Abstract

**Introduction:** Contrary to North America and Europe, the prevalence of hypertension is rising in West Africa. With a transition from whole foods to processed foods in Nigeria, diet plays a key driver of hypertension. To combat this, the national nutritional guidelines in Nigeria were implemented, but their translation into actionable tools for clinicians remains a challenge. Currently, there are no simple dietary assessment tools that are concise and suitable to be incorporated into clinical care without requiring extensive data analysis while still providing personalised dietary support to their patients. This study aims to deliver a clinically tested and validated short dietary assessment tool for clinicians, patients, and researchers across Nigeria to provide personalised dietary advice for patients with hypertension.

**Methods:** The study will be conducted in two phases: Phase 1 (n=75) will investigate the feasibility of the short FFQ and its agreement with 24-hour dietary recalls (3x) in a clinical setting in Nigeria. During the analysis of Phase 1 data, a scoring system will be developed based on the associations between individual food items in the FFQ and measures of hypertension. Phase 2 (n=50) will assess the acceptability of the FFQ and validate the association between the FFQ score and hypertension.

**Expected outcomes:** The development of a clinically tested and validated short food frequency questionnaire that will be ready to use by clinicians, patients, and researchers across Nigeria to support the prevention and management of hypertension.

**Conclusion:** This study will contribute to knowledge on dietary assessment and hypertension prevention by developing a validated and acceptable FFQ, which will be valuable for clinicians and researchers for personalised dietary recommendations to combat hypertension in Nigeria. ***Keywords:*** West Africa, diet, food, LMIC, blood pressure.

## 1. Introduction

### 1.1. Background and Rationale

Hypertension, defined as sustained high blood pressure, is the leading preventable risk factor for cardiovascular disease and the number one cause of death globally (1). Approximately 40% of people aged 30-79 years have hypertension, with two-thirds of cases living in low- and middle-income countries, including African countries (2, 3). The most recent data from the World Health Organisation (WHO) show that the African region has the highest prevalence of hypertension (35.5%), with the Americas having the lowest (18%) (4). Countries such as the UK, the US, and China have seen a decrease in the prevalence of hypertension, with rates declining by 6.25%, 11.38%, and 16.3%, respectively, from 2010 to 2019 (5–7). On the contrary, West African countries like Nigeria have shown a consistent increase in the prevalence of hypertension during this same time period. The most recent data estimate a 15.3% increase in hypertension rates in Nigeria from 2010 to 2019 [4,8], which has directly led to a documented increase in the prevalence of heart disease, stroke, and chronic kidney disease [9,10].

The increase in hypertension rates in West African countries has been attributed to unhealthy dietary practices and a lack of physical activity (1, 8). Among Nigerians, there has been a notable shift in dietary habits, with a significant rise in meals consumed outside the home and a growing preference for fast-food establishments offering processed foods high in fat, salt, and sugar (9). Unfortunately, this dietary transition has led to a surge in overweight, obesity, and the prevalence of diet-related non-communicable diseases, including hypertension, hypercholesterolemia, and diabetes (9). National nutritional guidelines were implemented in Nigeria in October 2014, providing evidence-based recommendations to reduce the risk of NCDs such as hypertension, diabetes, and obesity (10). However, the translation of these guidelines into actionable advice for clinicians for combating hypertension has been a challenge, possibly because (i) clinicians in Nigeria have not been provided with an effective strategy to provide regionally-specific dietary information to their patients, and (ii) the evidence used to inform Nigeria nutritional recommendations is based on evidence weighted towards non-Nigerian studies which may not be translatable or applicable to manage the contribution of diet towards the rising trend of hypertension risk in Nigeria and other West African countries.

Dietary assessment methods such as diet records and 24-hour dietary recalls are commonly used to evaluate an individual’s typical food and nutrient intake (11). However, these methods can be time-consuming and impractical in clinical settings (12). Alternatively, a brief food frequency questionnaire (FFQ) offers a more convenient and cost-effective approach to assess individual usual dietary intake and adherence to specific dietary patterns (12, 13). Nonetheless, effective dietary counselling requires a patient-centred approach that demands both time and expertise (14), making it challenging for clinicians and other health professionals to implement the use of dietary assessment tools in clinical care. Currently, there are no simple dietary assessment tools suitable for use by clinicians and healthcare professionals in clinical practice in Nigeria that are concise enough to be incorporated into clinical care without requiring extensive data analysis while still providing personalised dietary support to their patients. To overcome this gap, the study has set the following objectives: (1) to investigate the feasibility of a dietary assessment tool (FFQ) and its agreement with 24-hour dietary recalls (24HR) in a clinical setting in Nigeria and to develop a scoring system for the FFQ, and (2) to evaluate the acceptability of the FFQ and the association between the FFQ score and hypertension risk in Nigeria.

### 1.2. Study Aims and Objectives

#### 1.2.1. Study Aims

The overall aim is to deliver a clinically tested and validated short dietary assessment tool that will be ready to use by clinicians, patients, and researchers across Nigeria to provide personalised dietary advice to patients with hypertension and empower its citizens to improve their diet and take an active role in hypertension prevention.

#### 1.2.2. Study Objectives

1. To assess the feasibility of implementing the FFQ in a clinical setting in Nigeria.
2. To examine the agreement between the FFQ and 24-hour dietary recalls in capturing dietary information.
3. To develop a scoring system for the FFQ based on the associations between individual foods and continuous measures of hypertension (Blood pressure).
4. To evaluate the acceptability of the FFQ among the study participants.
5. To assess the association between the FFQ score and hypertension.

### 1.3. Trial design

This study will adopt a monocentric, retrospective, cross-sectional, controlled, open-label, nonrandomised design with parallel branches that will be conducted in a clinical setting at Rivers State University Teaching Hospital, Port Harcourt, Rivers State, Nigeria. The trial will adhere to the SPIRIT guidelines for effective scheduling and time management of trial participants **(Figure 1).** The study will be conducted in two distinct phases. Phase 1 will investigate the feasibility of the FFQ and its agreement with 24-hour dietary recalls, while Phase 2 will assess the acceptability of the FFQ and validate the association between the FFQ score and hypertension (**Figure 2**). Ethical approval has been granted by the University of Leeds, UK (AREA FREC 2023-0484-572) and Rivers State University Teaching Hospital, Nigeria (RSUTH/REC/2023316).

**Figure 1:**
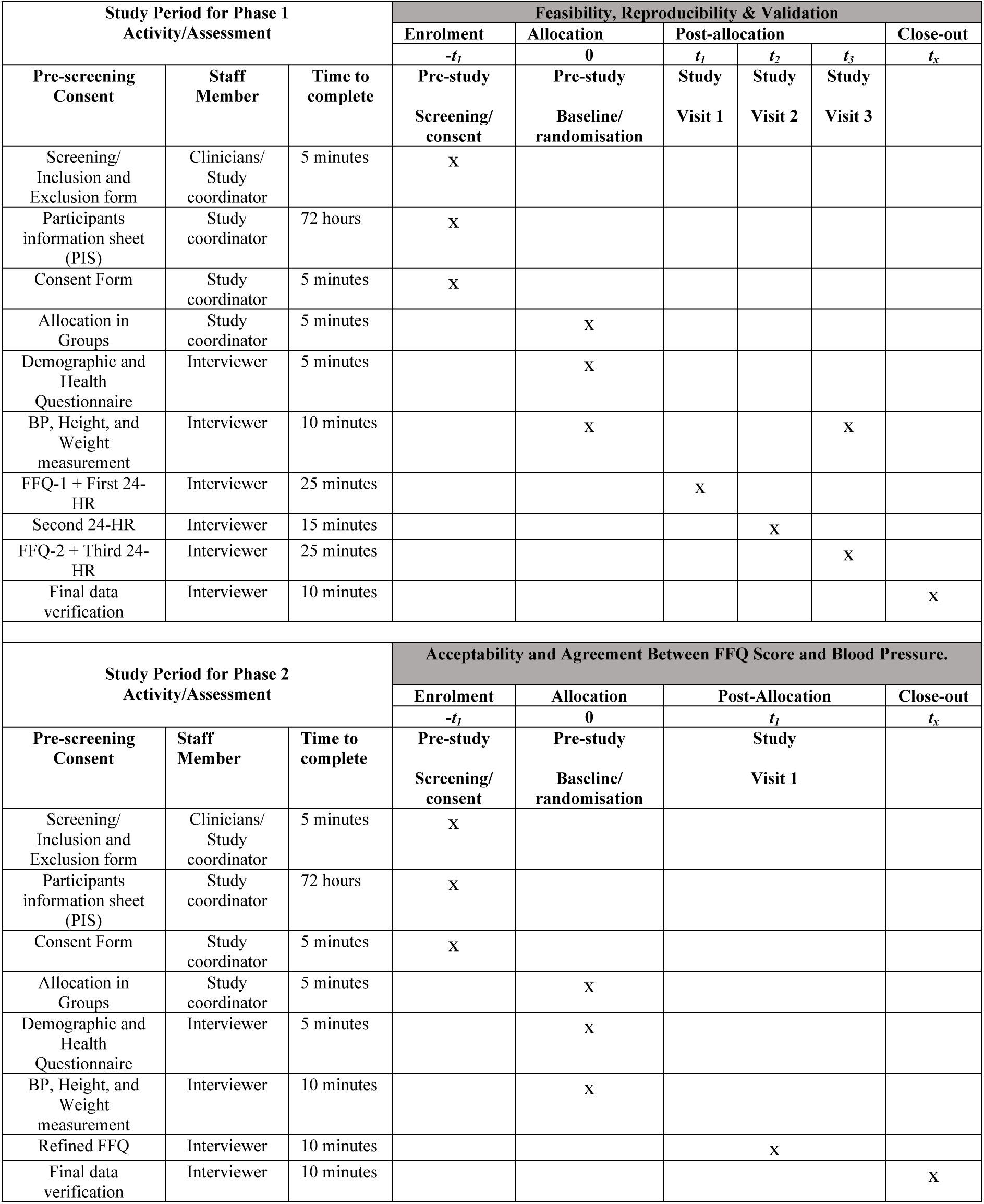
Schedule of enrolment, interventions, and assessments in accordance with the SPIRIT guidelines. t1: week 1; t2: between week 1and 3, t3: Week 3, t_x_: post-week 3.

**Figure 2.**
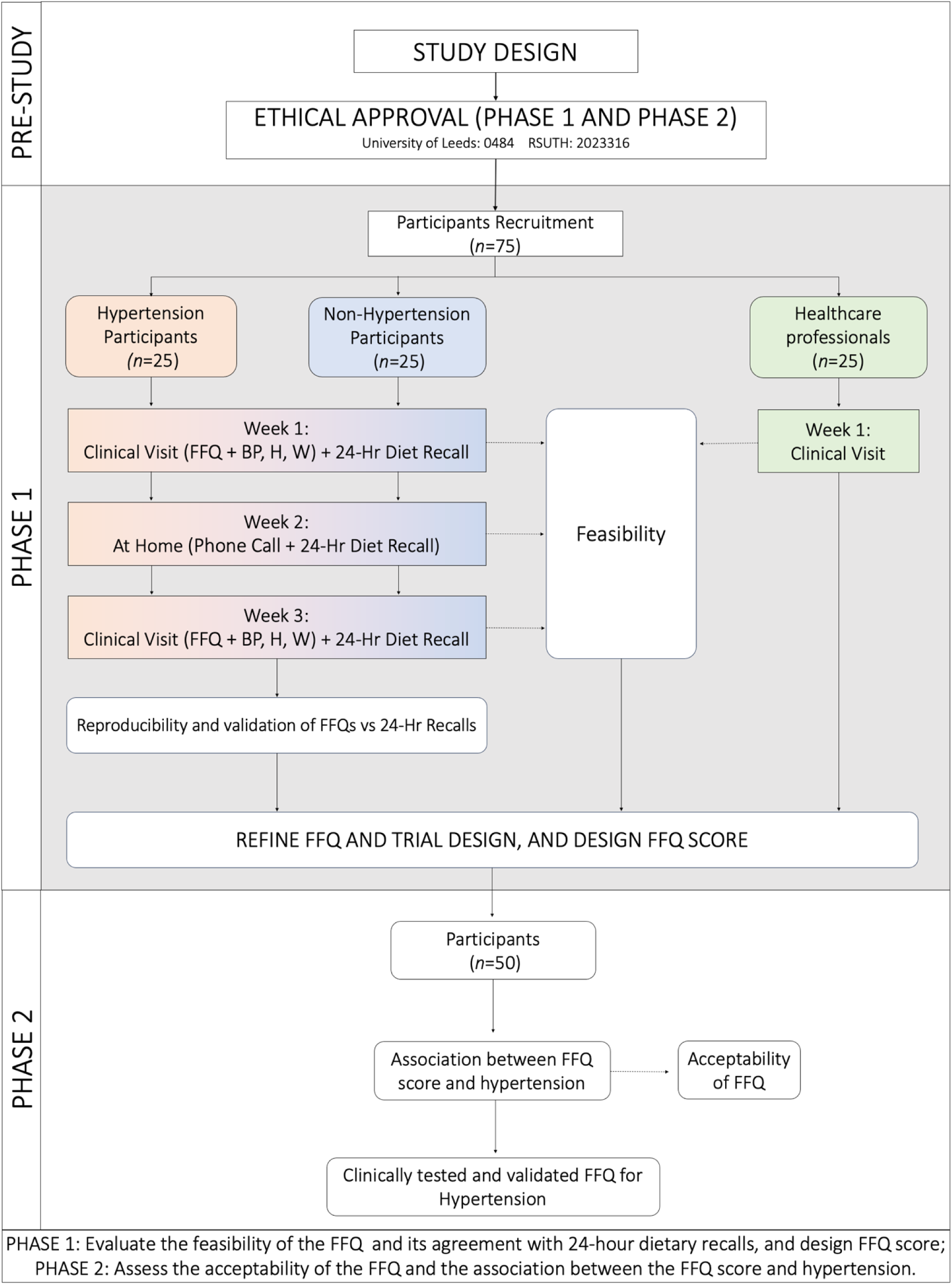
Participant selection and sequence of assessments flowchart. FFQ: food frequency questionnaire, 24 HR: 24-hour dietary recalls, BP: Blood pressure, H: Height, W: Weight.

## 2. Methods

### 2.1. Participants, Interventions and Outcomes

#### 2.1.1. Study setting

The study will take place at the Rivers State University Teaching Hospital in Port Harcourt, Nigeria. This clinical setting was chosen due to its accessibility to patients and well-established medical care and research infrastructure.

#### 2.1.2. Eligibility criteria

Interested individuals will be screened for eligibility during their regular clinical appointments, where clinicians, nurses, and the research team will use a short questionnaire to evaluate if they meet the inclusion of the study (**Table 1**).

**Table 1:** Eligibility criteria for the recruitment of study participants (clinical patients)

| Inclusion criteria | Exclusion criteria |
| --- | --- |
| Age between 18 and 70 years<br>Men and women<br>Hypertensive or non-hypertensive individual<br>Individuals resident in Nigeria for the past 2 years.<br>Ability to read, write, and communicate over the phone in English<br>Individuals who gave their consent to participate. | Individuals < 18 years or > 70 years of age<br>Pregnant women or intent to become pregnant<br>Breastfeeding woman<br>Diagnosis of other chronic diseases such as cancer, diabetes, renal failure, or endocrine diseases, and previous and recent cardiovascular disease (CVD) and stroke.<br>Individuals on dietary restriction or recent changes to their diet or food<br>Individuals who are currently enrolled in other studies |

#### 2.1.3. Intervention

##### Food Frequency Questionnaire (FFQ)

Food frequency data will be collected using the pre-printed self-administered food frequency questionnaire of 27 main food groups. For each food item, participants will be asked to report how many times they consumed the food over the past week (times/week). The FFQ will be self-administered to the participants in the clinics during their clinic appointment on the first and third visits.

##### 24-Hour dietary recall (24HR)

A 24-hour dietary recall (24HR) will be used to capture the habitual consumption of foods among the participants (15) and used as the reference method to validate the FFQ. The 24HR will be used to collect dietary information of the participants in the past 24 hours using the multiple-pass method (16, 17).

#### 2.1.4. Outcomes

##### Primary Outcome Measures

The primary outcome of this study is to deliver a clinically tested and validated short dietary assessment tool (FFQ) that will be shared with Nigeran Health Authorities and experts for implementation into Nigerian clinical practice. While this tool alone will not eliminate unhealthy diet and hypertension risk, it will be a major step towards positioning ‘diet’ as a practical means of hypertension prevention in Nigeria. This analysis of the result of the study will include descriptive data related to assessing the feasibility, validity, and reproducibility of the FFQ designed for the dietary evaluation and prevention of hypertension in Nigeria. On assessing the feasibility of the FFQ, gauging participants’ perceptions of clarity, ease of use, cultural appropriateness, and encountered difficulties while completing the questionnaire.

##### Secondary Outcome Measures

Secondary outcomes will evaluate the agreement between FFQ and 24-hour dietary recalls (24HR) in assessing dietary intake, using correlation coefficients to establish consistency between methods. Additionally, we seek to explore the association between FFQ scores and hypertension risk, quantifying the odds ratio that reflects the odds of hypertension about the FFQ scores. Furthermore, we will assess the acceptability of the FFQ through participant-reported feedback, providing insights into the practicality and user-friendliness of the tool.

#### 2.1.5. Participant timeline

The participant timeline and schedule of enrolment, interventions, assessments, and visits for participants are outlined in **Figure 1** in accordance with SPIRIT guidelines. During Phase 1, a one-month recruitment period will be initiated to identify clinical patients meeting the eligibility criteria. These patients will be approached for informed consent and subsequently categorised into two groups based on their history of hypertension. Participants will undergo comprehensive baseline assessments in the initial week of their clinical appointments. These assessments will encompass the administration of the Food Frequency Questionnaire (FFQ) and the First 24-hour dietary recall (24HR). The participant’s blood pressure and essential anthropometric measurements, including height and weight, will also be recorded. Moreover, demographic and health status information will be collected to overview the participants’ profiles comprehensively. A second 24-hour dietary recall assessment will be conducted in the subsequent weeks, followed by a final set of assessments in the third week post-allocation. These final assessments will include the administration of the second FFQ, the third 24-hour dietary recall (24HR), and recording blood pressure, height, and weight measurements.

Phase 2 of the study will involve participants who have provided informed consent. Over four weeks, participants will undergo assessments during their clinical appointments. Central to this phase is the refined FFQ-based dietary intake evaluation, designed to capture detailed information on participants’ dietary habits. Simultaneously, height and weight measurements will be taken to ensure accurate and up-to-date anthropometric data. Additionally, relevant demographic and health status information will continue to be collected, allowing for a comprehensive understanding of participants’ backgrounds and overall health profiles. This phase provides a detailed and in-depth analysis of participants’ dietary habits and their potential correlation with hypertension status.

#### 2.1.6. Sample size and sampling technique

A non-probability convenience sampling method will be used to recruit eligible participants from the outpatient clinics at Rivers State University Teaching Hospital (RSUTH). Previous studies examining the correlation between food frequency questionnaire (FFQ) and 24-hour dietary recall have found a strong agreement with correlation coefficients (*r*) ranging from 0.3-0.7 (18–21). Based on this, a modest *r* = 0.5 is considered a good indicator for determining the sample size needed to evaluate the agreement between our clinical FFQ and 24-hour recall (22). For phases 1 and 2 of the study, a statistical power of P = 0.8, a 95% confidence interval, and a two-tailed alpha level of 0.05 were chosen. Using G*Power software (23) to estimate the required sample size based on the correlation coefficient (*r*=0.5), it was determined that a minimum sample size of 29 participants would be needed. In addition, recent evidence suggests that 13 participants are adequate to achieve sufficient qualitative feedback (i.e., saturation) in studies with a small number of clear objectives (*See Section 3.4.2: Feasibility assessment of FFQ*) (24). As such, we will also be sufficiently powered to evaluate the qualitative evidence and feedback with 29 participants. Nonetheless, to account for an anticipated dropout rate of 20% and to accommodate any potential missing/incomplete data, the target sample size will be set to 50 participants (**Figure 2**) (25, 26). Therefore, a sample size of 50 participants (clinical patients) will be deemed sufficient to achieve saturation for the feasibility, validation, and acceptability goals in phases 1 and 2 of the study. In addition, we will aim to recruit 25 clinical healthcare professionals (i.e., nurses and clinicians) at RSUTH to provide qualitative feedback on the questionnaire.

#### 2.1.7. Recruitment

The study participants will be adult clinical patients and will be recruited from the Rivers State University Teaching Hospital (RSUTH) outpatient clinics in Port Harcourt, Rivers State, Nigeria. The recruitment methods for phases 1 and 2 of this study will involve a multifaceted approach. Participants (clinical patients) will be recruited through recruitment posters displayed in key hospital areas, clinician referrals, and engaging discussions during morning briefing sessions where vital signs are taken from clinical patients in internal medicine and family medicine departments. The healthcare professionals will identify eligible participants based on study criteria, and informational posters will provide essential study details and refer interested participants to the study team for further screening.

### 2.2. Assignment of Interventions

Participants in this study will be allocated into non-hypertensive and hypertensive groups based on their history of hypertension, as illustrated in **Figure 2**. Allocation will be conducted following predefined criteria and without randomisation. The implementation of group allocation will be carried out by the primary investigator based on the predetermined criteria. Participants will not be informed of their assigned group during their respective clinic appointments. Allocation will be conducted in a standardised and systematic manner to uphold the integrity of the study.

### 2.3. Data Collection, Management and Analysis

#### 2.3.1. Data collection

##### PHASE 1

###### Feasibility assessment of FFQ

The feasibility of the FFQ will be evaluated in a small number of participants (n=75) in a ratio of 2:1 consisting of 50 clinical patients and 25 healthcare professionals to ensure adequate saturation in both groups (**Figure 1**). In the feasibility assessment, eligible clinical patients and healthcare professionals will fill out the questionnaire and be asked to provide feedback on the clarity, ease of use, cultural appropriateness and difficulty encountered in answering the questions. Additionally, the healthcare professionals will provide feedback on the clinical relevance and potential impact on patient care and suggest improvements to optimise the questionnaire’s effectiveness. In summary, we will be well-powered to evaluate feasibility from the perspective of patients and clinicians.

###### Validation and Reproducibility of the FFQ

The FFQ will be administered to clinical patients (*n*=50) twice (i.e., first and third visits) to quantify the intake of food items. Additionally, an inconsecutive three (3) repeated 24HR, including two weekdays and one weekend day, will be used to collect dietary information from the clinical patients for the past 24 hours using the multiple-pass method (16, 17) at the time of visits and intervals between the two FFQs administrations (**Figure 1**). The reproducibility of the FFQ will be assessed by measuring the level of agreement between the two administrations of the FFQ. Furthermore, the validation will be assessed by evaluating the agreement level between the FFQ and the average of the three 24-hour recalls (27, 28). Healthcare professionals are not involved in the patient FFQ validation stage.

###### Development of FFQ score

The FFQ score will be developed using the participant’s dietary intake captured by the FFQ (**Figure 1**). A three-point scoring system will be developed for each of the food groups - healthy dietary intake (3 points), moderate dietary intake (2 points) and unhealthy dietary habits (1 point) – with participants scored according to their self-reported intake (29, 30). The scoring system will be informed according to existing scientific evidence, dietary guidelines and recommendations, and validation studies (31).

##### PHASE 2

###### Acceptability assessment of the FFQ

A feasibility trial to test the acceptability of the refined FFQ and gather information on modifications to improve ease of use and future implementation in Nigerian clinics will be conducted (**Figure 1**). The FFQ will be administered to clinical patients (n=50) once to collect their dietary intake and feedback to assess the acceptability of the FFQ. Acceptability will be evaluated by qualitative feedback on indicators of the (i) process (e.g., proportion interested, started, completed FFQ), (ii) resource (e.g., time to complete and review, rated performance on handheld devices), and (iii) data management (e.g., suitability of user interface, data storage and analyses) endpoints(25). Ten questions, 5-item Likert-scale questions, such as “strongly disagree, disagree, neutral, agree, or strongly agree”. The ratings for each of the 10 questions will be used to generate both individual participants’ acceptability ratings as well as an overall acceptability score from all participants.

###### Assessment of the FFQ score and hypertension risk

The refined FFQ will be administered to the clinical patients (n=50) to assess their dietary habits. Individual FFQ scores will be calculated for each clinical patient from their FFQs. A logistic regression will be performed between FFQ scores and hypertension risk (blood pressure), with a correlation coefficient (r) of around 0.5, suggesting a moderate to strong association. The odds ratio will be calculated to quantify the strength of the association between the FFQ score and hypertension.

##### Non-Dietary Data Collection

In phases 1 and 2, in addition to dietary intake, socioeconomic and demographic data such as gender (sex), age, education, and level of physical activity will be collected using a structured questionnaire from the clinical patients. The blood pressure and anthropometric data (such as weight and height) will be measured using a standard procedure from each clinical patient, and the average will be calculated and used. Body mass index (BMI) will be calculated using weight in kilograms by height in meters squared. Hypertension will be defined as SBP ≥ 140mm Hg and DBP ≥ 90mm Hg or on antihypertension medication (32).

#### 2.3.2. Data Management

Data management, confidentiality, and access in this study will adhere to stringent security measures. All data collected in physical format (such as Questionnaires, including the primary eligibility screening questionnaire, health questionnaires, and Food Frequency Questionnaires (FFQs)) utilised during participant visits will be digitised through scanning. Both electronic and physical copies will be retained, with the latter securely stored within a locked cabinet in the principal investigator’s secure office.

#### 2.3.3. Statistical Analysis

Statistical analyses will be conducted using R programming language version 4.3.0 (33) with a significance level set at p<0.05. Descriptive statistics will be used to analyse categorical and continuous variables. Categorical demographic variables will be analysed using frequency distribution, while continuous demographic variables will be summarised as mean and standard deviation for normally distributed variables and median and interquartile range (IQR) for non-normally distributed variables. As the frequencies of food groups are not normally distributed, differences in frequencies between the two FFQs (FFQ1 vs. FFQ2) and between the two methods (24 HR vs. FFQ2) in phase 1 will be tested using Wilcoxon’s signed-rank test.

Feasibility and acceptability will be assessed through descriptive analyses, including means and 95% confidence intervals (CI), qualitative feedback, and content analysis of indicators related to the process (e.g., proportion interested, started, completed FFQ) and resource (e.g., time to complete and review, rated performance, and practicality).

The West African Food Composition Table will be used to calculate the nutrient content (macronutrients such as energy, carbohydrates or sugar, proteins, fats, and fibre and minerals such as sodium and potassium) of the foods reported in both the FFQ and 24HR. The mean and distribution of nutrient intakes obtained from the FFQ and 24HR will be calculated, and a comparison will be made to assess the differences using the Wilcoxon signed-rank test. Pearson correlations will be used to calculate the correlation coefficients between the FFQ and 24HR for each nutrient to assess the agreement between the FFQs and 24HR and the reproducibility of the FFQs. Additionally, the agreement between the FFQ and the average of three 24-hour dietary recalls will be estimated using Intraclass Correlation Coefficient (ICC) or quintile cross-classification analysis and the Bland-Altman plots will be used to visualise the agreement between the recalls and FFQ (34–36).

Dietary intake according to the 27-item FFQ will be used to develop an FFQ score based on associations between individual foods and blood pressure using correlation and regression analysis. The result of the regression models will inform the weighting and scoring assignment for each food item in the FFQ. Following the assignment of scores for each individual food item, an aggregation process will be undertaken to obtain an overall dietary score for each participant. The association between the FFQ score and blood pressure will be evaluated using regression analysis, allowing for a robust evaluation of the scoring system’s predictive validity and adjusting for potential confounders, such as age, sex, BMI, physical activity, and socioeconomic status.

### 2.4. Monitoring

This study will implement a robust data monitoring plan to ensure the collected data’s accuracy, integrity, and confidentiality. Trained research personnel will regularly review and verify the data for completeness and consistency. Additionally, electronic data capture systems will facilitate real-time monitoring and error detection. Any discrepancies or anomalies will be promptly addressed and documented.

Given the non-interventional nature of this study and the utilisation of non-invasive data collection methods such as the 24-hour dietary recall, the anticipated risks and harms to participants are minimal. However, participants will be informed of any potential discomfort or inconvenience associated with data collection.

Additionally, measures will be in place to address any unexpected adverse events that may arise during the study. To ensure adherence to established protocols and regulatory guidelines, periodic audits will be conducted by an independent third party. These audits will encompass a comprehensive review of study documentation, data collection procedures, and adherence to ethical standards. Any identified discrepancies or deviations will be addressed promptly, and corrective actions will be implemented to maintain the integrity and validity of the study.

### 2.5. Ethics and Dissemination

#### 2.5.1. Research Ethics Approval

Ethical approval has been carefully addressed in this study, as evidenced by the approval from the following ethics boards. Business, Earth & Environment, Social Sciences (AREA FREC) Committee, University of Leeds, Leeds, United Kingdom, approved with the number: 0484 on 28/04/2023. Similarly, the Rivers State University Teaching Hospital Research Ethics Committee in Port Harcourt, Nigeria, approved with the number: RSUTH/REC/2023316 on 30/03/2023. The study prioritises the protection of participants’ rights and well-being by upholding key ethical principles. All participants will undergo the informed consent procedure to ensure that all participants will receive comprehensive information about the study’s purpose, methods, potential risks, and benefits. All data collected, including questionnaires and personal identifiers, will be treated as confidential. Participants will be assigned unique study IDs, and strict security measures will be implemented.

#### 2.5.2. Protocol Amendments

Any proposed amendments to the protocol that could impact the conduct of the study, participant recruitment, participant information dissemination, or data analysis will undergo a thorough evaluation. These modifications will be submitted as amendments to the University of Leeds Research Ethics Committee for approval. Following their approval, the updated protocol details will also be communicated to *ClinicalTrials.gov*. In cases where changes pertain to enrolled participants, they will be promptly informed of the modifications.

#### 2.5.3. Consent

Participants who meet the inclusion criteria will be given the Participant Information Sheet (PIS) and an informed consent form to review independently. After 3 days, study personnel will contact the eligible participants by telephone. If they express interest in participating in the study, they will be asked to sign the informed consent and be given a study identification number.

#### 2.5.4. Confidentiality and Access to Data

All data collected will be coded to ensure anonymity, and any identifying information will be securely stored separately from the research data. Throughout data collection, a stringent anonymisation process will be implemented. Participants’ identities will be safeguarded by recording only their randomisation numbers and dates of birth within the data collection tools. Anonymised data will be treated in accordance with the University of Leeds’s Research Data Management Policy, ensuring that sharing, publication, and utilisation are conducted while preserving participant privacy. Any personal, identifiable, or sensitive information will be excluded from publications. Data retention will extend for a minimum of 5 years following publication. Access to the data will be restricted to authorised research team members exclusively.

#### 2.5.6. Dissemination Plans

The results from this study will be disseminated promptly through various channels to ensure wide accessibility and impact. First, a manuscript detailing the research methodology, results, and conclusions in both phase 1 and phase 2 will be prepared for submission for publication in open-access, peer-reviewed scientific journals. Secondly, presentations will be made at relevant national and international scientific conferences, allowing for engagement with experts in the field and the broader research community. In addition, the results will be summarised in accessible and easy-to-use formats for dissemination to healthcare professionals, policymakers, and the public in Nigeria. Infographics, fact sheets, and webinars will be developed to convey key findings and practical implications for hypertension prevention strategies. The entire protocol, the anonymised dataset and any relevant statistical code will be deposited in the Leeds Research Data Repository and available open access.

### 2.6. Discussion

Unhealthy dietary shifts from whole foods to highly processed ‘fast food’ options have been linked to the escalating hypertension prevalence in Nigeria. Notably, dietary habits are evolving, marked by increased consumption of meals from eateries and fast-food vendors offering high-fat, high-salt, and high-sugar processed foods (1, 8, 9). Positive outcomes in hypertension and cardiovascular health have been noted through dietary and lifestyle changes (37–40). Hence, responsive short dietary assessment tools can swiftly guide healthcare professionals in pinpointing concerns and establishing and tracking dietary goals for patients, including patients with hypertension (41, 42). Several tools exist for dietary management of cardiovascular disease, type 2 diabetes, hypertension, and non-alcoholic fatty liver disease (NAFLD) that have successfully been developed for clinical use (27, 41, 42). Nevertheless, many of these tools have been designed and validated primarily in the US and Western European nations, limiting their applicability to Nigeria. Our primary aim is to explore the feasibility, validation, and acceptability of a short dietary assessment tool for evaluating dietary intake among individuals with hypertension in Nigeria.

Currently, no short dietary assessment tool responsive to dietary changes has been developed for hypertensive patients and incorporated into clinical care for the prevention and management of patients with hypertension in Nigeria. Given the practicality of FFQs in settings lacking formal dietetic support and their inherent inclusion of portion sizes to minimise measurement error [15], our aim is to validate a developed short FFQ. The developed FFQ will be informed by the findings of Batubo et al. (8) and the Nigeria Nutritional Guidelines (10) culminated in a 27-item tool designed for use with hypertensive patients. We aim to investigate the relative validity of our newly developed short FFQ in relation to 24-hour dietary recall from 50 clinical patients focusing on commonly consumed food in Nigeria. This approach to validate the FFQ has been extensively utilised in the literature by previous studies (27, 29, 43–45).

Feedback from clinical patients and healthcare professionals regarding the FFQ’s feasibility is expected to drive its refinement within a user-friendly clinical context. This could potentially bridge the gap between nutritional guidelines and clinical care, equipping healthcare professionals to systematically assess the dietary intake of patients for diagnostic and preventive purposes and enable clinicians to provide region-specific dietary recommendations, fostering improved disease prevention and management, including hypertension (46). The anticipated findings from the Intraclass Correlation Coefficient (ICC) analysis and correlations between the FFQ and 24-hour dietary recall (24HR) are poised to illuminate the FFQ’s reproducibility and validity in assessing dietary intake. Strong ICC values and correlations above 0.4 would indicate the FFQ’s reliability and accuracy in capturing habitual dietary patterns, which is crucial for providing accurate dietary advice to patients (42, 47, 48). The scoring system for the FFQ could offer a practical means of categorising habitual dietary habits. Should the scoring system align with established dietary recommendations and scientific evidence, it could be a valuable tool for clinicians to assess dietary quality and offer tailored dietary guidance to patients. The assessment of the acceptability of the FFQ. Positive feedback could indicate that the FFQ is well-received by clinical patients and that its implementation could be feasible in a clinical setting. This information is essential for potentially integrating the FFQ into routine clinical practice.

This study protocol presents several notable strengths. Firstly, the study addresses a critical gap in the rising challenge of hypertension in Nigeria and the lack of a short dietary tool that can be incorporated into clinical care by focusing on the feasibility, validity, and acceptability of a dietary assessment tool explicitly tailored to Nigeria. Secondly, the inclusion of both clinical patients and healthcare professionals in the feasibility assessment phase ensures comprehensive feedback from both user groups, enhancing the applicability and relevance of the dietary assessment tool. The limitations of this work include the use of a sole patient cohort and the risk of selection bias, which means that the results may not be generalised to other populations or settings beyond the selected clinical outpatient. Furthermore, reliance on self-reported dietary data through FFQs and 24-hour recalls introduces potential recall bias and measurement error, which may impact the accuracy of dietary assessments (49, 50).

## Data Availability

No datasets were generated or analysed during the current study. All relevant data from this study will be made available upon study completion.

## Acknowledgements

Not applicable

## Supporting Information

SPIRIT Checklist for the Protocol

Informed consent material

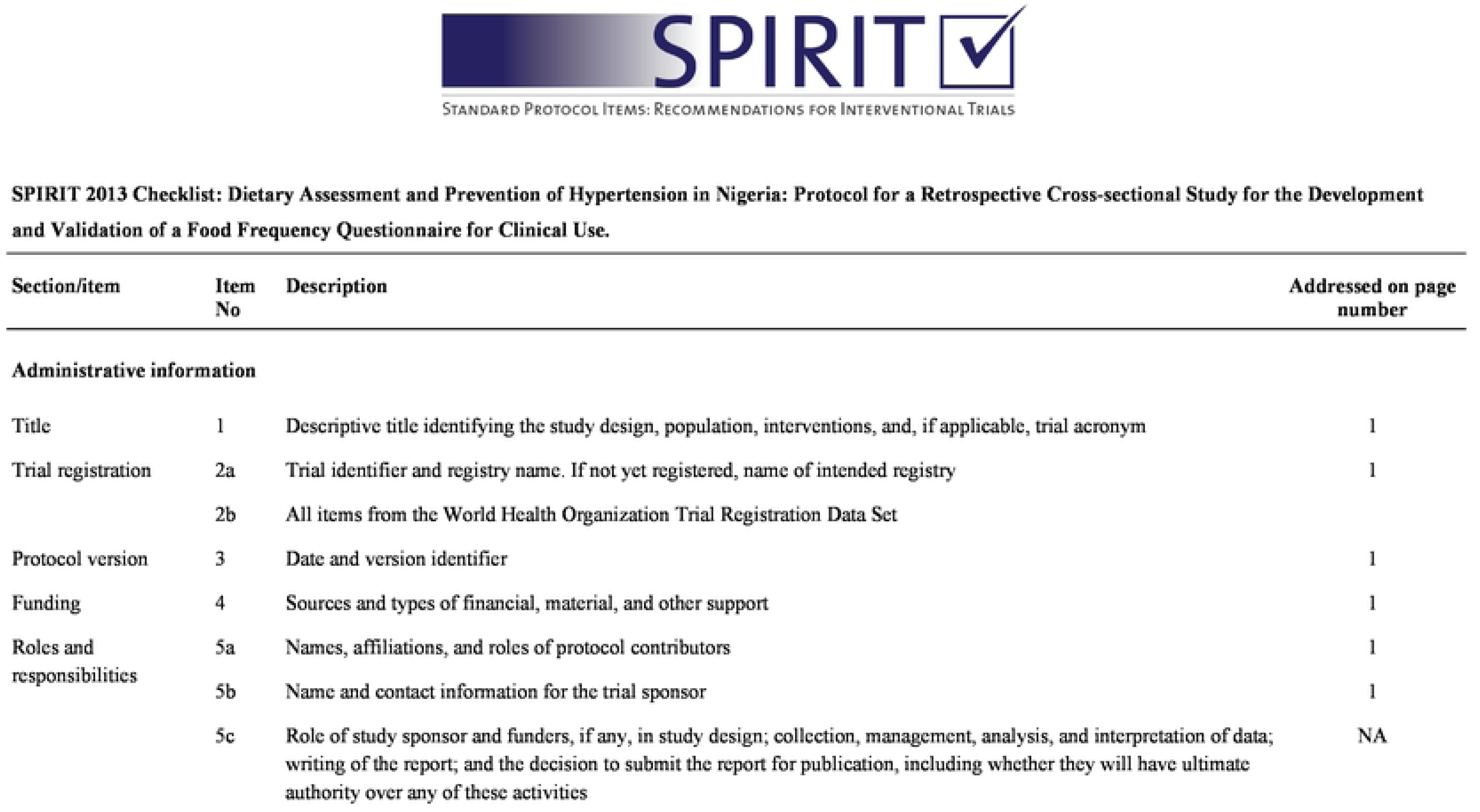
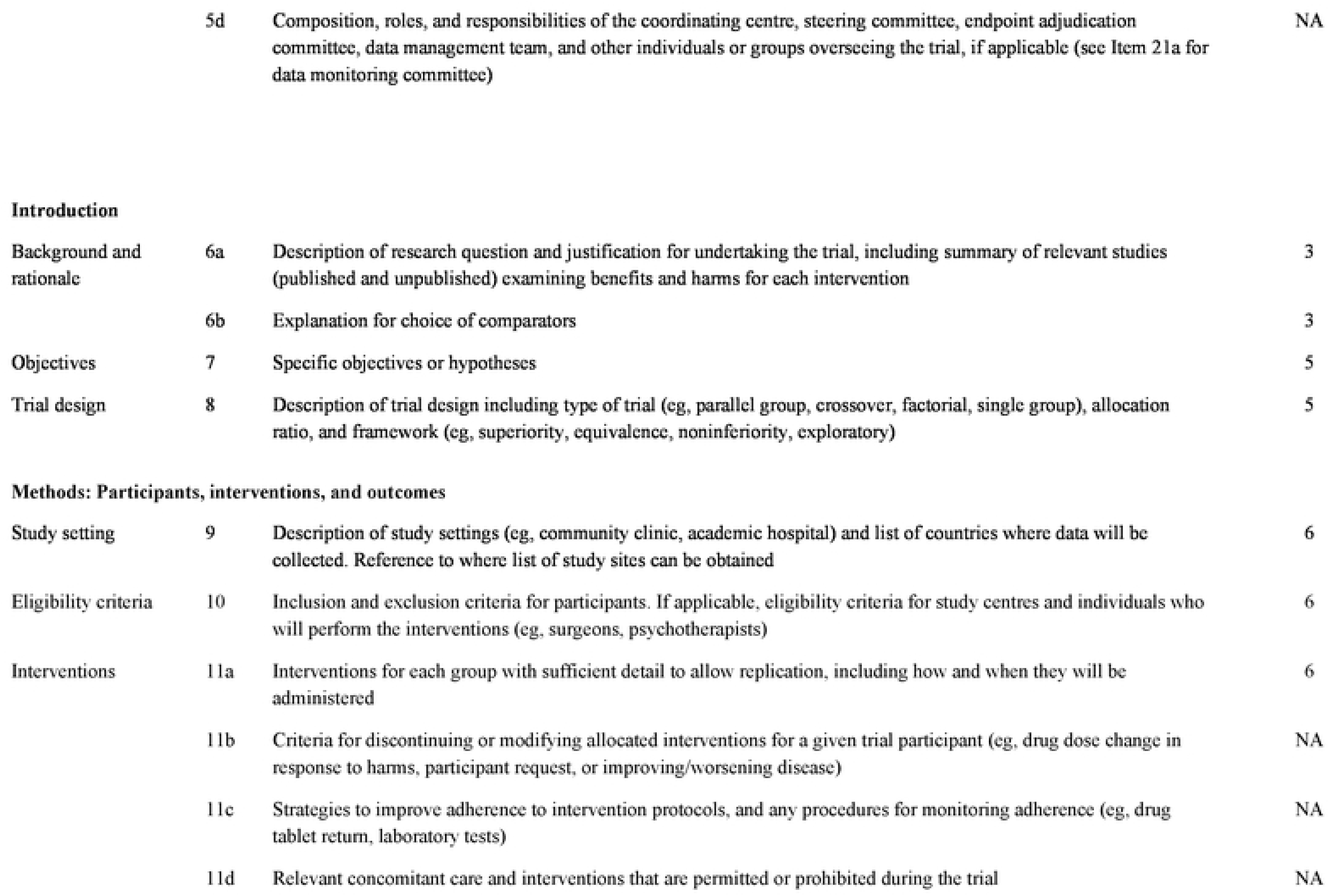
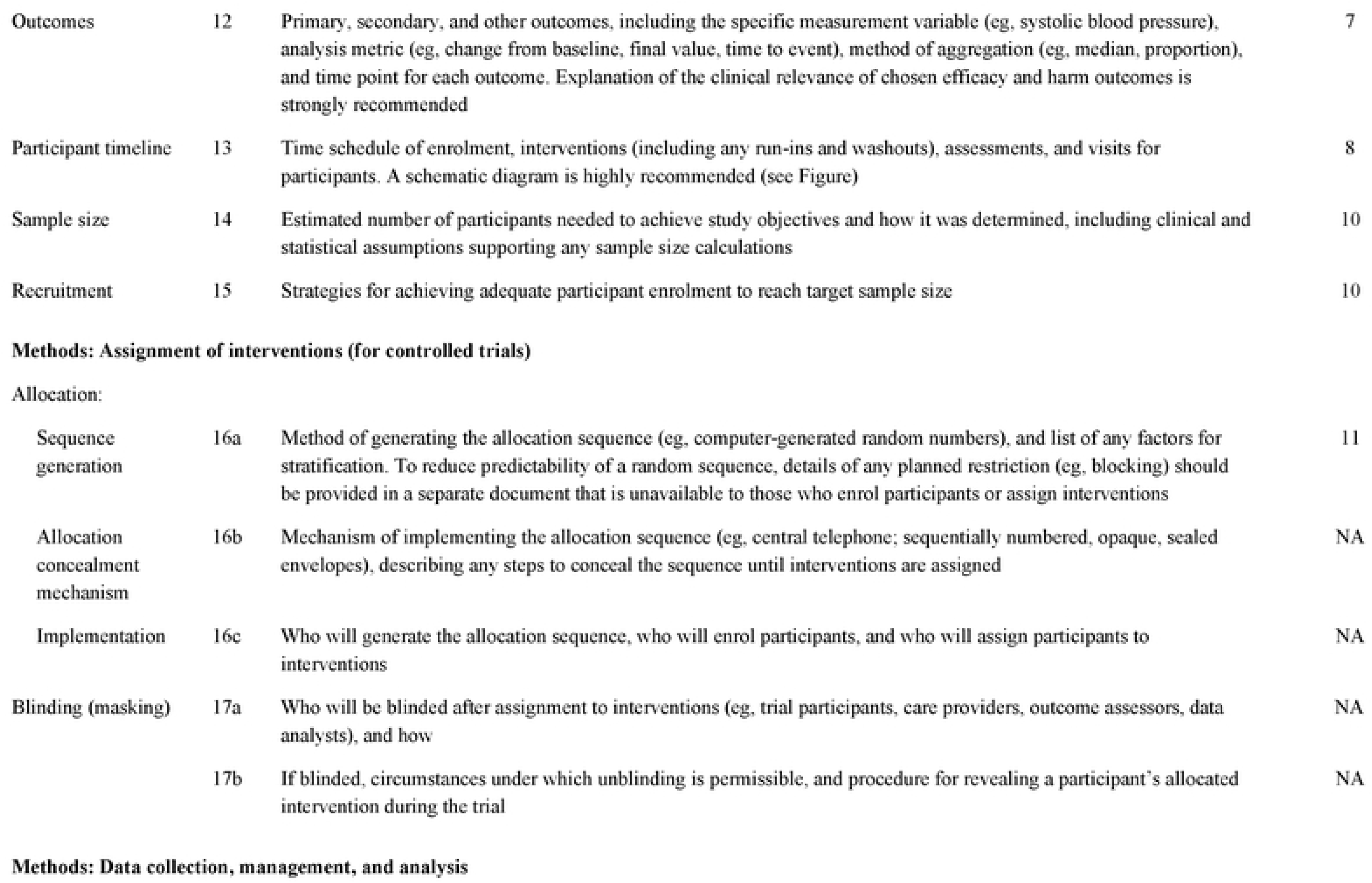
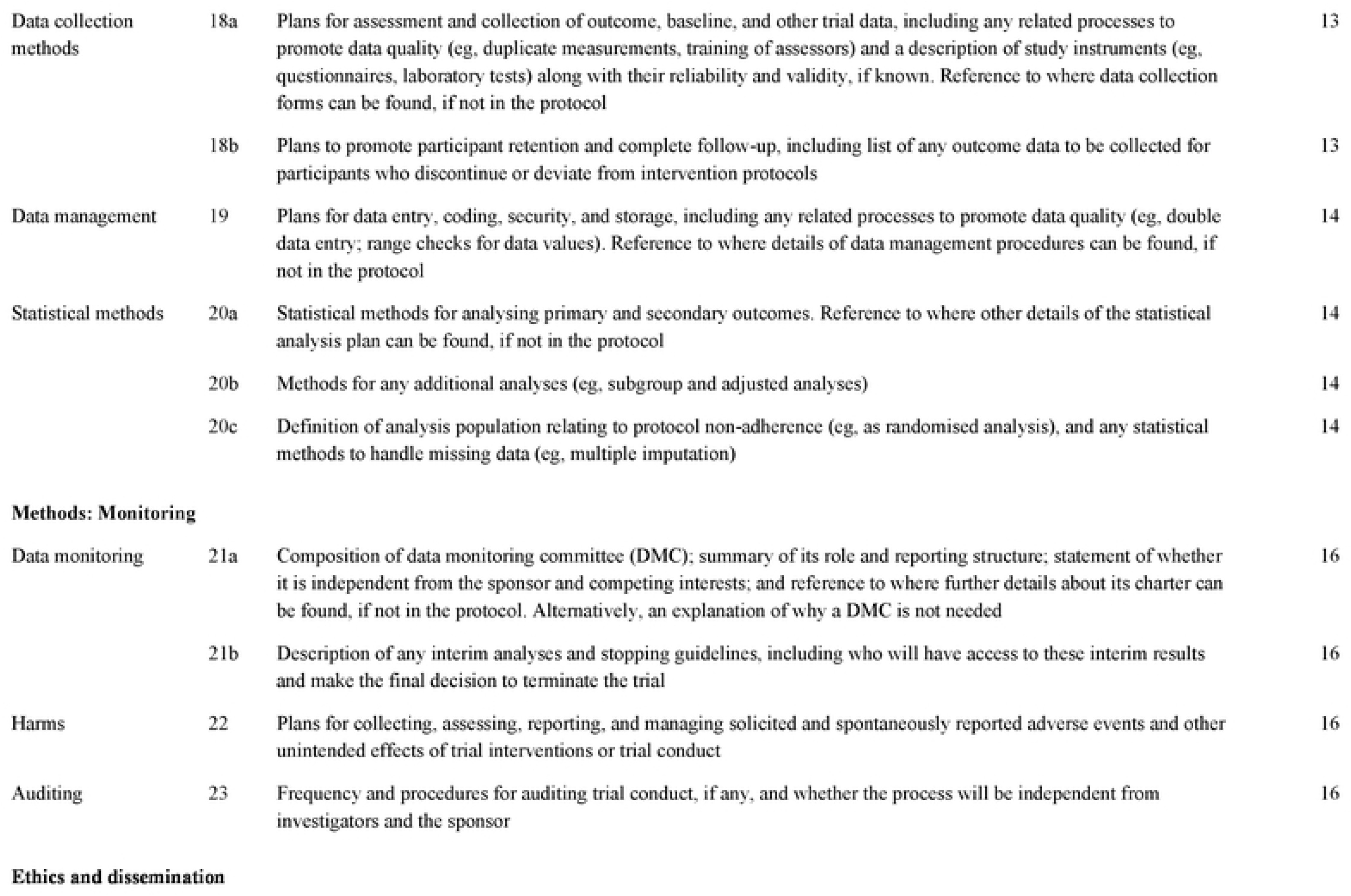
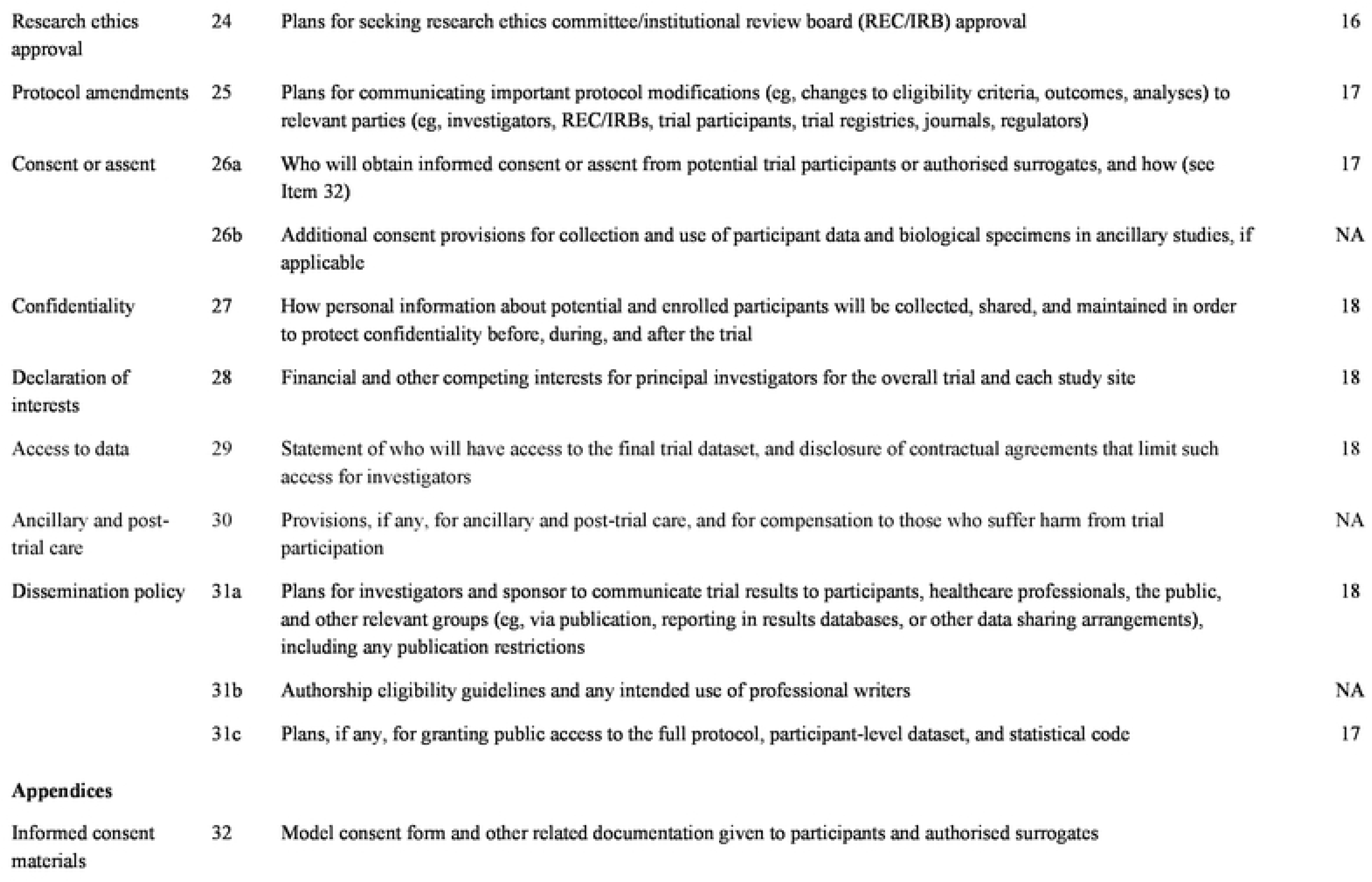
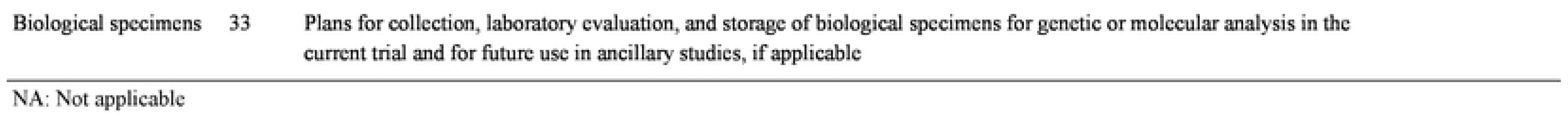

